# COVID19 is a seasonal climate-driven disease across both hemispheres

**DOI:** 10.1101/2020.12.16.20248310

**Authors:** Alejandro Fontal, Menno J. Bouma, Adrià San José, Mercedes Pascual, Xavier Rodó

## Abstract

The role of climate in the population dynamics of COVID-19 remains poorly understood, and a true seasonal signature has remained elusive. Data from both hemispheres and the second wave provide opportunities to further examine climatic drivers. With a statistical method designed to detect transitory associations, we show consistent negative effects of temperature and absolute humidity at large spatial scales. At finer spatial resolutions we substantiate these connections during the seasonal rise and fall of COVID-19. Strong disease responses are identified between 12-18°C for Temperature and 4-12 g/m^3^ for Absolute Humidity. These results classify COVID-19 as a seasonal low-temperature infection, and point to the airborne pathway as an important contribution to transmission for SARS-CoV-2, with implications for control measures we discuss.

## Introduction

With the periodic influenza pandemics in the 20th century, the Spanish flu in 1918, the Asian flu in 1957, Hong Kong flu in 1968, the medical community has been apprehensive about the emergence of novel pathogens with an extensive reach in a more connected world (1). Apart from influenza with its well appreciated ability to mutate (2), the few relative benign corona viruses have not caused alarm. That was until the 2003 outbreak of Severe Acute Respiratory Syndrome, SARS-CoV-1 (SARS) in China (3), and Middle East Respiratory Syndrome (MERS) in Saudi Arabia in 2012 (4) with severe symptoms and high mortality. Both outbreaks were short-lived and isolation of cases proved sufficient to overcome the restricted ability of these pathogens to transmit between humans. The emergence of the novel SARS-CoV-2 with a similar geographic and zoonotic origin as SARS (5) was declared a pandemic by WHO in February (6) and has rapidly spread over extensive regions, with morbidity often overwhelming regional healthcare capacity. So far, over 63 million cases have been recorded, with over 1,4M deaths (7), and a high mortality, not unlike influenza, in the older age groups (8).

The initial propagation of COVID-19 emerged in a latitudinal band between 30°N and 50°N with low humidity levels and temperatures between 5°C and 11°C (9), and these weather sensitivities were reported from China, with absolute humidity (AH) negatively associated with daily death counts of COVID-19 (10). SARS-CoV-2 further resembled influenza with respect to its winter appearance. These initial observations suggested SARS-CoV-2 could be a seasonal disease, a possibility that was not advanced for the short-lived outbreaks of SARS and MERS. Recent perspectives have reviewed and evaluated the evidence for climate-driven seasonality, indicating contradictory results and confounding effects of other factors at these early stages of the pandemic (11). Mathematical modelling further addressed the potential for seasonality in the population dynamics of COVID-19 by parameterizing a humidity-dependent transmission rate on the basis of seasonal influenza models and time series data for other coronaviruses (12). Results indicated an inability of climate forcing to establish seasonality given the large number of available susceptible, non-immune, individuals fueling transmission.

At a more mechanistic level, some observations on long range transmission of SARS in 2003 (13), raised the scope for aerial transmission and a role of aerosols (3), which would bypass the short-range control measures that dominate current control response. The intangible connection between airborne transmission and seasonal transmission has been implicit in the scientific interest that followed the SARS 2003 outbreak, particularly for influenza. Both the nature of seasonality and the plausible significant contribution of airborne transmission were explored (14–16) and attempts were made to mechanistically connect temperature, humidity to environmental persistence of bio-aerosols and seasonality. Strikingly, the results of these influenza studies on aerosols are similar to those of initial SARS-CoV-2 investigations (17), implying a possible contribution of aerosols to long range transmission. With the second pandemic wave now underway in the Northern Hemisphere, the question of COVID-19 being a genuine seasonal disease is acute, with implications for what would be judicial intervention measures.

This second wave makes now possible the further analysis of climate drivers, in a way that takes advantage of consistent effects across countries and regions, and the rising and waning phases of the epidemic. Further weight to pattern analyses is given here by consistency of results across temporal and spatial scales, with a statistical method for transient correlations that identifies similar variation in time series over localized windows of time.

## Results

To address the association of temperature (T) and absolute humidity (AH) on the initial phase of the invasion by SARS-Cov2 and before changes in human behavior and public health policies came into place, we first considered a measure of the initial growth rate of cases across countries. For this, we relied on an estimate of the reproductive number of the disease, *R*_0_, in the 20 days after the first 20 cases of COVID-19 were officially notified for each of 162 countries spanning 5 continents (Fig.1A, Methods, SI). The global maps for the climate variables at those corresponding times in each country and for population density are shown in Fig. S1.

**Fig. 1.**
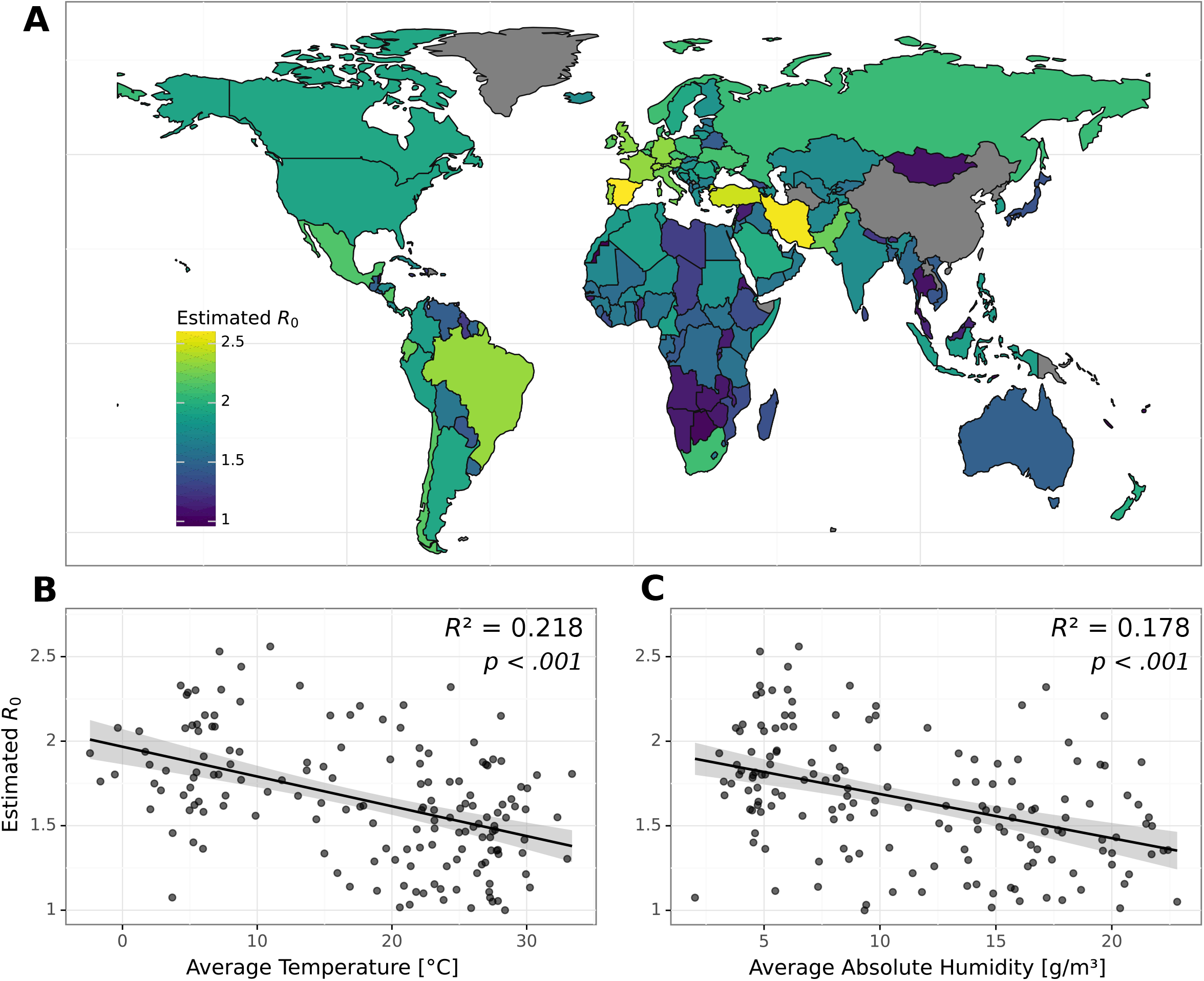
Estimated basic reproduction number (*R*_0_) during the initial phases of the pandemic for all countries included in the study (n=162). (A) To focus on the initial rise of the first wave, these estimates are based on the cumulative COVID-19 cases during the first 20 days following the 20th confirmed case in each country (see Methods). Scatterplots showing the variation of the estimated R0 for the different countries as a function of their average temperatures [°C] (B) and absolute humidities [g/m^3^] (C) during the initial phase of the pandemic. Black lines and *R*^2^ values correspond to the estimates of an univariate linear model in both cases, fitted by Ordinary Least Squares (p<0.001 in both cases). Of the 162 countries included globally, the numbers corresponding to each continent are the following: Africa (n=50), Asia & Oceania (n=45), Europe (n=38), North America (n=16), and South America (n=12). For comparison, the *R*_0_ estimates obtained after the first 100 reported cases (rather than 20) are shown in Fig. S1. Similar spatial patterns are observed. The spatial distribution of average temperatures and absolute humidities, as well as the national weights based on relative population density, used to compute these averages, are shown in Fig S1(B-D).

A negative relationship is observed between *R*_0_ and T, as well as AH, at the global scale on the basis of linear pairwise regression models (Fig.1B,C and Fig. S1). Although these regressions account for 10%-25% of the variance in *R*_0_, the two regressions are consistent and statistically significant (p<0.001), irrespective of the COVID-19 measure employed (e.g. *R*_0_ or cumulative cases; see also sensitivity tests in Fig. S2).

To address whether the negative relationship with both T and AH is merely the result of a coincident timing between trends in epidemic spread and wintertime conditions in temperate regions, we analysed the evolution of the disease-climate associations locally in time and at several disaggregated spatial scales for groups of countries in all five continents (Fig. 2). To this end, we relied on a statistical method, Scale-Dependent Correlation Analysis or SDC, specifically developed to identify similar patterns of variation by means of local or transitory (linear) correlations between two time series given a moving window of time (Methods, and (18–20)). Application of this analysis to other diseases known to be climate sensitive has successfully shown that (linear) associations can be discontinuous in time, alternating temporal intervals with high significant correlations and those with low or non-existent ones(21, 22). Such transient coupling can be expected in the response of nonlinear systems to an external driver, especially when the functions relating particular parameters to the driver are themselves nonlinear, so that effects are more evident in particular ranges of the forcing variable. SDC is essentially a pattern-recognition tool which allows one to consider the scale over which to evaluate correlations, as the window size can be systematically varied to focus for example on increasingly local patterns of variation.

**Fig. 2.**
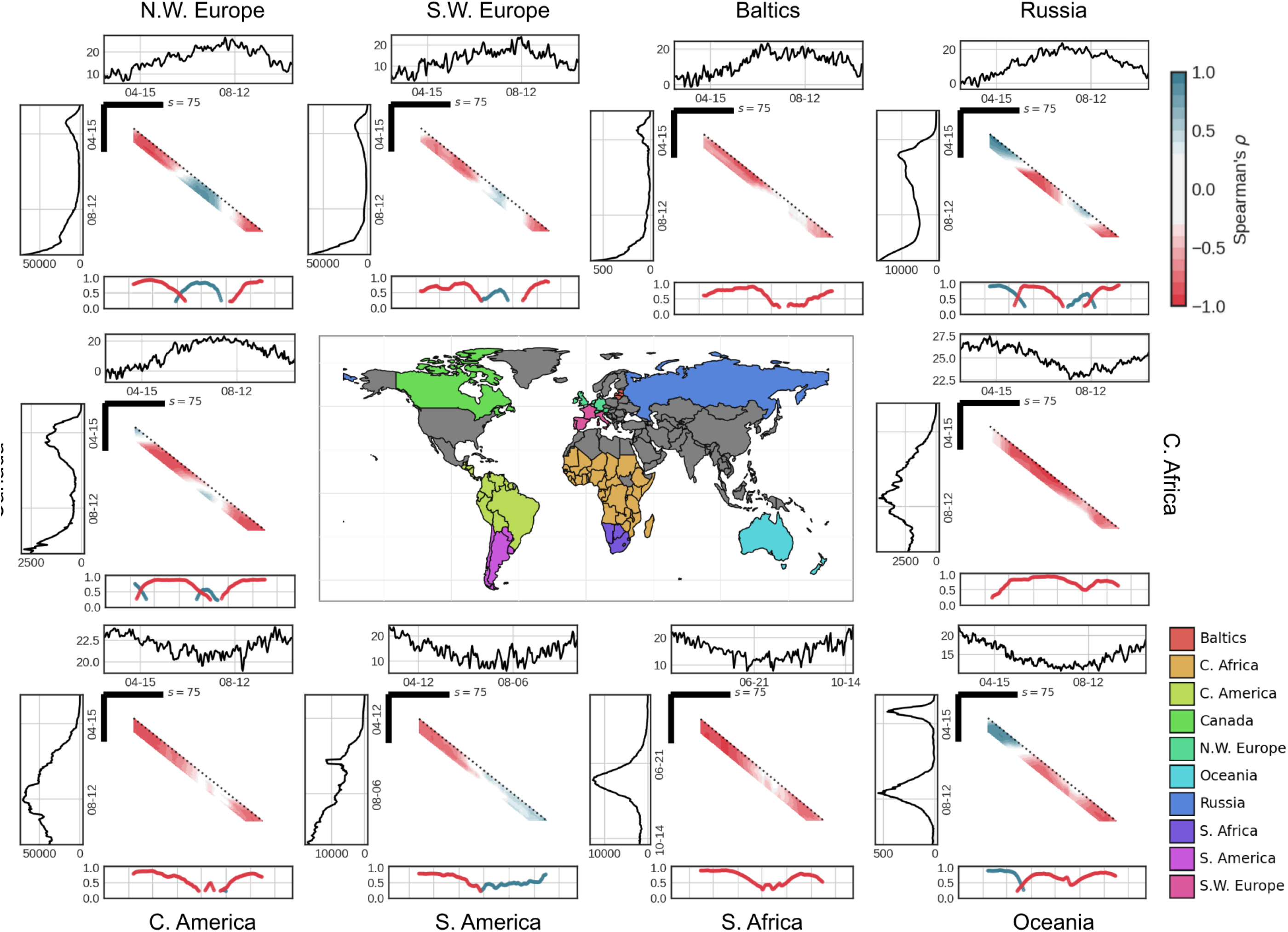
Two-way Scale-Dependent Correlation (SDC) analyses for the time series of COVID19 daily new cases vs. mean temperature (A) in groups of contiguous countries. These daily climate conditions are weighted averages based on the relative population of the relevant countries from March to October 2020. The temporal scale or window size s used for the local correlations is 75 days, and the lag between the placement of the windows in the two time series varies from 0 to 21 days. In the SDC plots, the two time series are shown respectively to the left and top of the central correlation grid, for reported cases with time running down and for the climate factor with time running right. Each cell of the grid is coloured according to the Spearman correlation coefficient, with the row and column corresponding to the position of the two respective time windows s along each of the time series, with the lag between these position corresponding to the distances from the diagonal (lag 0 to +21 days). The panel below shows the maximal correlation coefficient obtained at each time point (vertically, and therefore, relative to the time of the climate time series). Only correlations found as significant in a non-parametric randomized test (α=0.05) are shown and colored. Red (blue) points in the grid correspond to negative (positive) correlations, according to the specified scale (top right).

Figure 2A shows the results of SDC for COVID-19 cases and T for time windows of two months and a half (e.g. scale of s=75 days) when grouping countries according to region and latitude in the two hemispheres. Similar patterns are found for cases and AH (Fig. S3). Strong negative transient associations are obtained for short time lags between the disease and climate time series, with consistent patterns worldwide. Interestingly, the negative relationship is seen during both the first and second waves of the pandemic, and for both the rising and declining phases, with a break during summertime in all continents (Fig.2 and Fig. S3). Transient positive correlations of varying intensity are also detected during the warmer months across location, which are later discussed. These vary in intensity and are not consistently high.

To further examine the association patterns, we next considered the reported cases at the smaller spatial scale of individual countries in Europe (Fig.3 for France, Great Britain, Italy, Spain and Germany), the first most affected continent following the emergence of the virus in China. SDC results show similar transient and negative associations for T and AH with COVID-19 cases (s=75 days). The negative relationship occurs largely in synchrony across the different countries, for the same time intervals during the waning of the first epidemic wave as T and AH rise, and also during the rise of the second wave in the Fall as T and AH fall, with a break in between (Fig.3). For this same temporal window (s=75), a similar negative relationship holds also for individual regions within three of these highly affected European countries for which data at the higher spatial resolution of individual provinces are available (namely, Lombardy, Thüringen and Catalonia, Fig. S4). These results are also shown separately for the first and second waves in Lombardy (Fig. S4A), Thüringen (Fig. S4B) and Catalonia in Spain (Fig. S5C). Locally in time, these associations account for large fractions of the variability in COVID-19 cases in the three regions (over 80% at scales of s=21 days). Their discontinuity explains the lower values obtained with the traditional correlation coefficient, since by definition this quantity averages over the whole length of the time series (20).

**Fig. 3.**
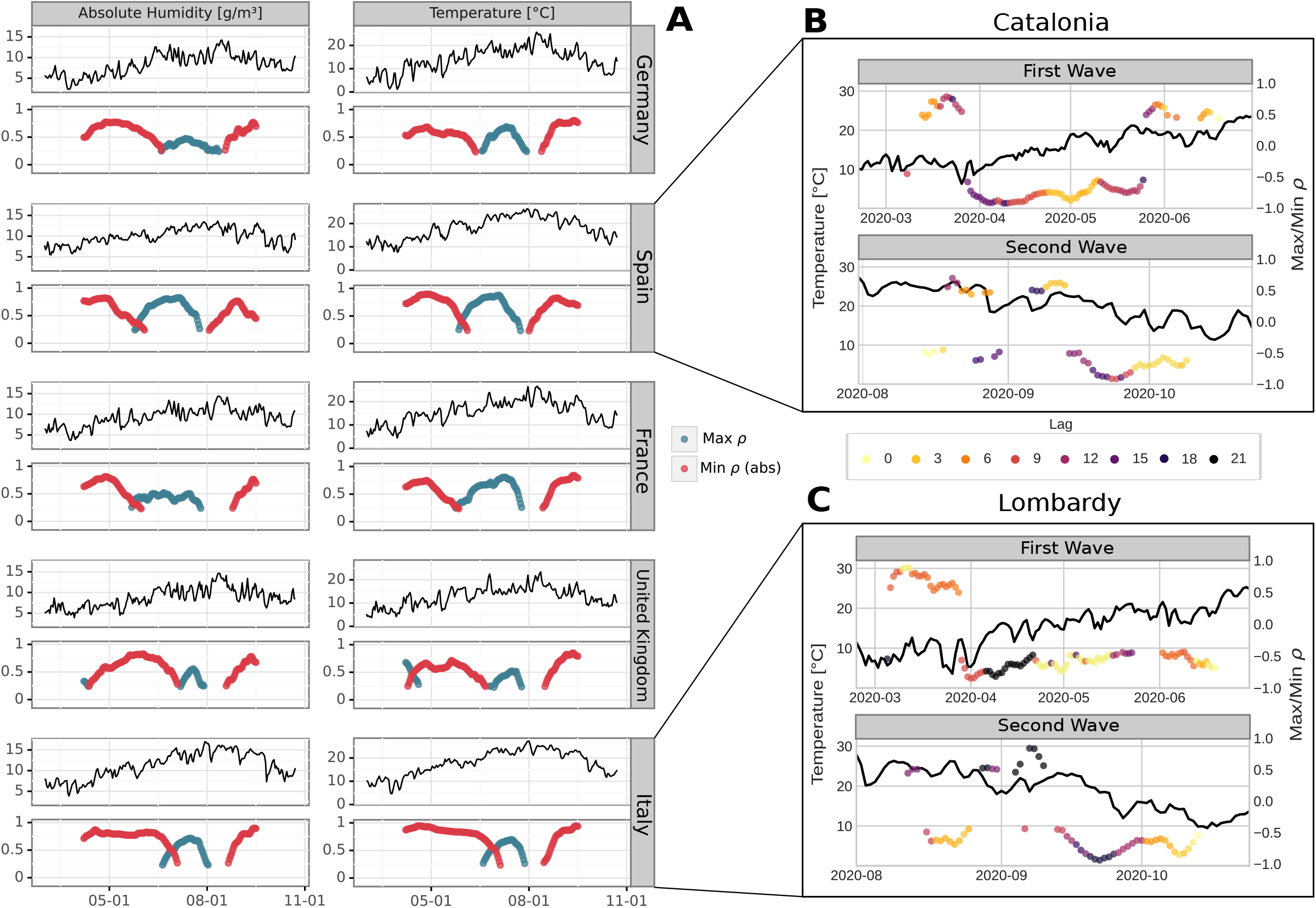
Panels from SDC analysis for the maximum absolute correlations between daily time series for the meteorological variable (AH, left; T, right) and COVID-19 new cases, as a function of time (the position of the local window of time). These panels correspond to those shown below the SDC grid of local correlations in Figure 2, but here at the country level for 5 European countries (Germany, France, Italy, Spain and UK). As before, s=75, and the maximum correlation is chosen for lags from 0 to 21 days. The colors correspond to maximum negative (red) and positive (blue) correlations for each day. Resulting maximum correlations for the Spanish region of Catalonia with the corresponding time lag indicated (B). This dominant lag (the one for the maximum ρ value among the significant comparisons between a lag 0 and +21 days) is overlaid on the temperature time series for each time point, with shorter lags being represented with lighter colours and longer lags with darker ones. The fragment size (s) used in this panel is 21 days, which allows us to localize and identify multiple shorter transient couplings between COVID19 incidence and meteorological factors. C) Idem as B) but for the Italian region of Lombardy. Similar figures for the German region of Thüringen can be found in Fig. S6. Note that in panels B and C when the meteorological factor appears to be more related to the transmission of SARS-CoV-2 and the spread of COVID-19 in the population, the lag shortens, indicating a more intense and fast association.

We can also inspect how these transient couplings change as a function of the temporal lag in days between the two time series. As the magnitude of the change in the climate variable T or AH increases, the lag tends to shorten (see boxes in Fig. 3B and Fig. S6). This behavior is seen for the direction of change that is presumably relevant for influencing transmission (increasing T and AH in the fall of the first wave, and decreasing T and AH in the rise of the second wave). The shorter lags may reflect a faster speed of community transmission, concomitant with a more intense effect of the meteorological factors.

Zooming in even more locally in time with a window of about two weeks (s=14), SDC reveals limited intervals of very high significant correlations at these regional levels for Lombardy (Italy; Fig.4) and for Catalonia, Spain (Fig. S7). Intervals where the climate covariate accounts for over 80-90% of the variability in cases alternate with complete decoupling between the time series. Interestingly, the strongest negative associations are observed during the on-going rise of the current second wave in these European regions. Positive associations are weaker and infrequent at this scale.

**Fig. 4.**
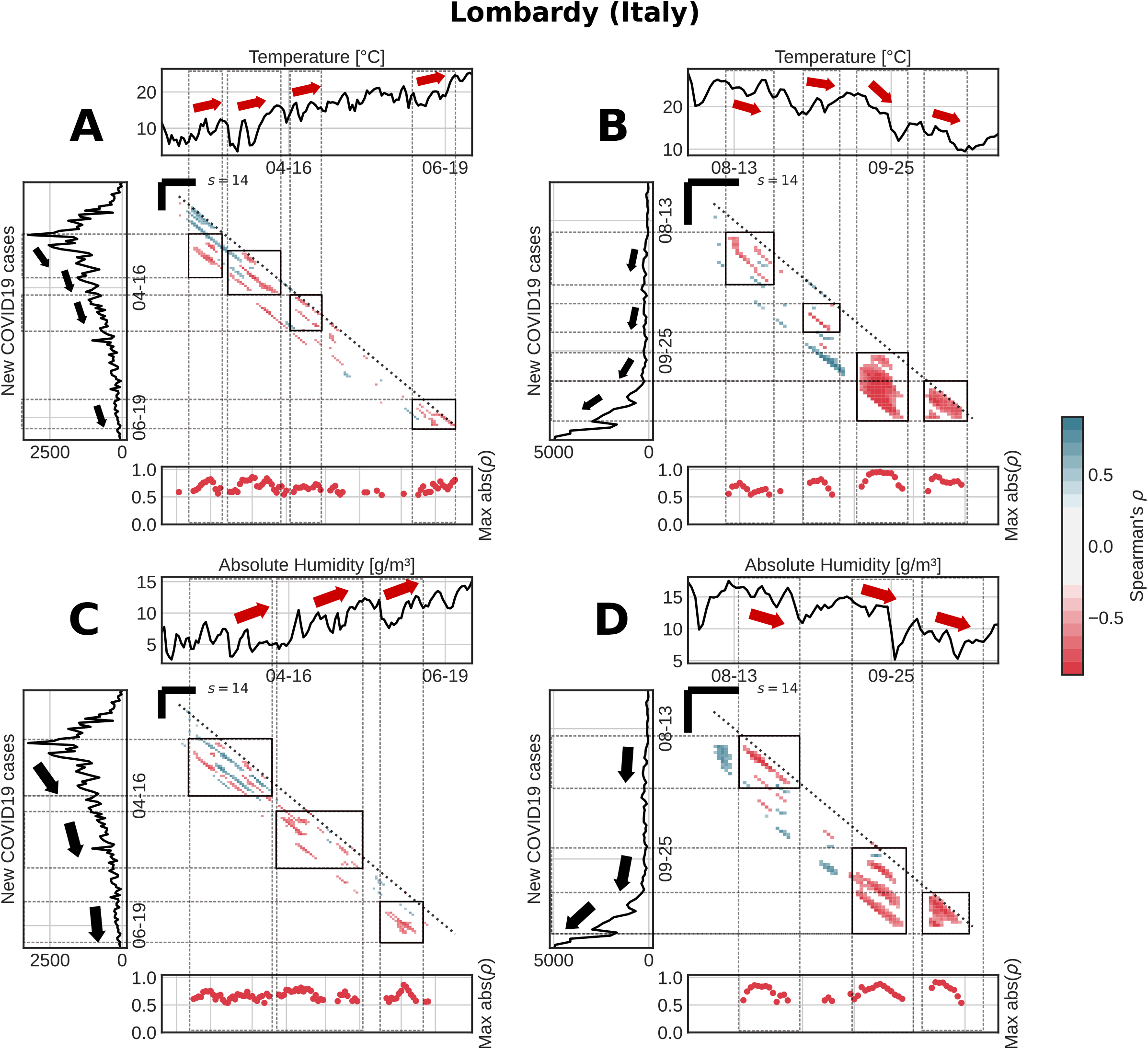
Short-term transient correlations over time intervals of two weeks at the spatial resolution of region, here for Lombardy in Italy. As for Figure 2, the graphical results of SDC analysis are shown for the correlations between daily Covid19 cases and T (top row) or AH (bottom row) and for the different parts of the pandemic. Here, a smaller window size is used (s=14 days) to examine even shorter-term transient associations. The left side panels (A, C) showcase the results for the waning phase of the first wave (from late February to July), and the right side panels (B, D), those for the rising phase of the second wave (from August to late October). The time intervals of significant correlations are indicated with boxes within the central grid, with the corresponding timings in the two time series highlighted with red arrows for the meteorological factor, and with black ones for the response variable, the COVID-19 cases. The pairs of arrows for a given box show clear patterns of opposite temporal change (negative correlation) in the climate and epidemiological variables. For clarity, in this case the bottom panels below each plot show only the maximum negative correlation values obtained as the analysis window is slided in time (with red dots indicating negative correlations).

Finally, we interrogate our results to address the existence of threshold values in T and AH for the spread of COVID-19 To this end, we examine whether intervals of coupling become more frequent depending on the range of the climate covariate. This behavior would indicate the existence of a critical range within which responses are heightened, and conversely outside which the forcing becomes ineffective. A meta-analysis was performed for Italy where detailed regional data on cases are available. We jointly analysed all SDC correlations obtained between COVID-19 cases and each of the climate covariates (T or AH), by pooling together the results for all the individual regions of Italy (Fig.5; Fig. S8; Methods). The resulting distributions in Fig. S8(A,B) show the proportion of the total possible comparisons (pairs of time intervals between the two time series) that fall into a given range of the climate covariate, with those proportions subdivided into significant positive and negative correlations (Fig.5A,B) and including non-significant ones (Fig. S8). Distributions are also shown for only the positive and negative correlations along the range of climate intervals (Fig. 5A,B), and for the two waves separately (Fig. S9, Fig. S10). The mode in these distributions indicates preferential intervals for T and AH respectively, where the climate effects are most evident and beyond which decoupling is likely (12-18 °C and 4-12 g/m^3^, respectively; Fig.5C-D). Similar results are obtained for the first and second waves separately (Fig. S10).

**Fig. 5.**
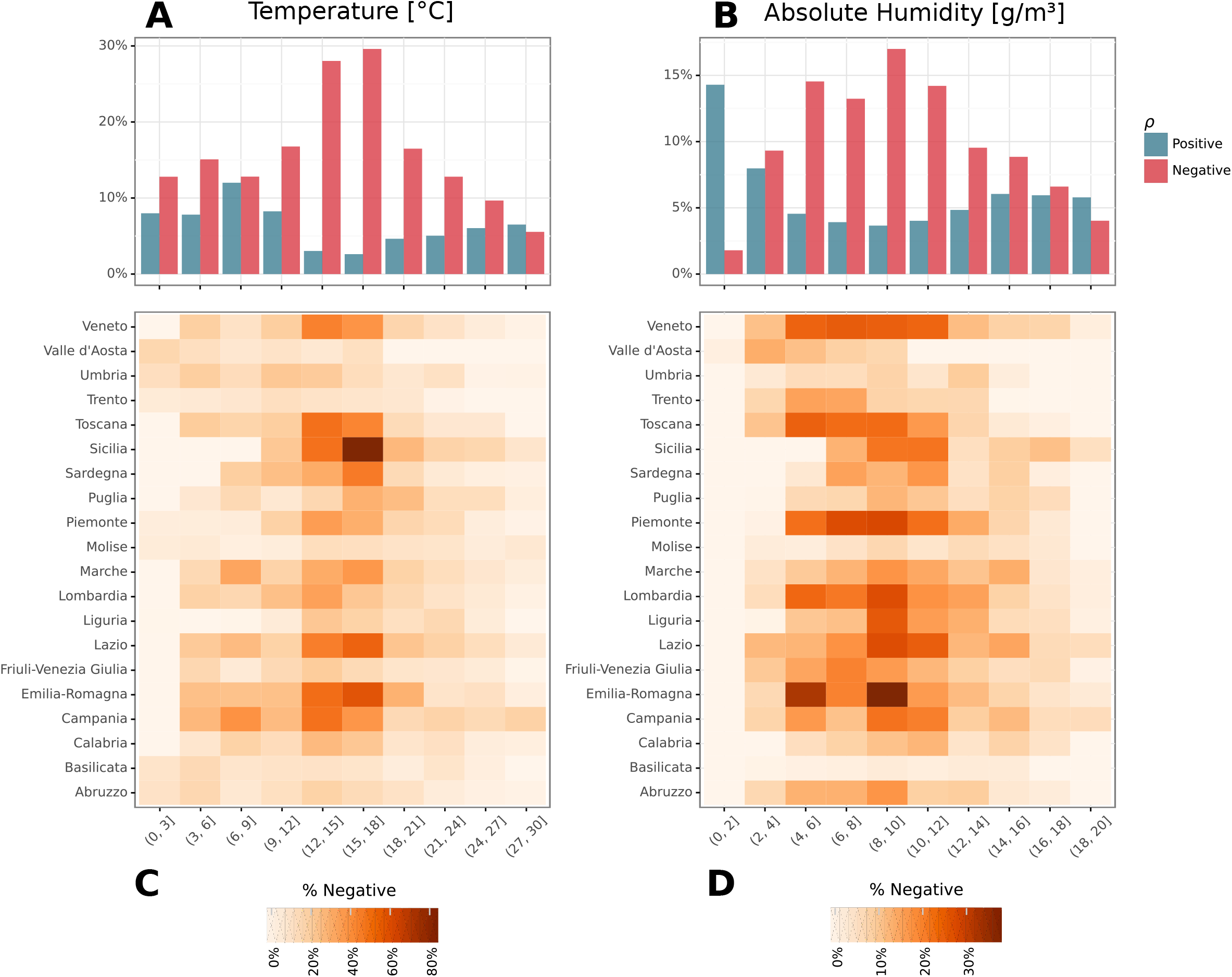
Frequency distribution of negative and positive correlations as a function of range of the climate covariate, for temperature (A) and absolute humidity (B). The frequency of windows with negative (or positive) correlations is obtained from SDC analysis of COVID19 cases and the given climate factor in all italian regions (n=20) (for s=21 days). The distributions are non-uniform with a mode in a particular range of the climate variables. This is indicative of a nonlinear relationship where the negative associations are most likely to be observed in a particular meteorological range. The bottom panels show this same frequency distribution but across the different italian regions (C,D). As it can be seen, there is an overrepresentation of significant negative monotonic relationships in temperate conditions (AH [4-12] g/m^3^, T [12-18] °C).

## Discussion

In summary, we observed consistent negative correlations between COVID-19 and weather conditions, T and AH, for regions and individual countries. This consistency across scales is observed for the rise of the first wave, its decline towards the advancing summer in both hemispheres, and the second wave after the summer. These associations remain robust and become stronger on finer temporal scales within countries during the seasonal rise and fall of cases, reflecting similar patterns of temporal variation between disease incidence and climate factors. The strong correlations become more frequent where and when seasonal mean temperatures fall between 12°C - 18°C, and AH levels, between 4g/m3-12g/m3, conditions that diverge from those observed for the mid-winter global emergence of SARS-CoV-2 (9), suggesting permissive temperatures above 5°C. Our results are also consistent with earlier studies more limited in scope (23–25). Taken together, the findings support the existence of strong external drivers of transmission intensity, as suggested by a uniform summer recession despite a variety of intervention measures across countries (26). They also support the view of COVID-19 as a true seasonal infection, similar in that sense to the more benign circulating corona viruses (27). Concomitant temporal variation of ecological conditions, human behaviour and disease incidence over the annual cycle can produce true but also spurious correlations with climate parameters (19). Our analyses were formulated to circumvent the spurious correlations arising from such coincident seasonal variation, as well as effects of potential confounders. The global correlation between Ro and climate covariates across countries specifically considered the timing of initiation of the first epidemic in each location to avoid the confounding effects of later interventions. Then, by addressing correlation patterns locally in time and zooming in for finer scales, the SDC analysis detected time intervals with similar patterns of variation that are not simply the result of ups and downs over the seasons or the typical durations of intervention policies. Identification of significant correlations between temporal patterns at different and embedded window lengths is considered an efficient way to infer plausible causality with this multiscale approach (18, 28). The implemented randomizations indicated that the spurious occurrence of the described correlations from noisy fluctuations is highly unlikely (e.g. see Fig. S11). Together with the above-mentioned consistency, these methodological considerations add weight to the findings.

The problem of co-linearity in climate factors has challenged the search for environmental drivers of influenza in the past, and very similar issues now apply to COVID-19. Both T and AH appear here equally capable of accounting for the observed COVID-19 variation. It is well-known that rising air-temperature can contain more water, and for geophysical reasons these two parameters are usually strongly coupled over extensive spatial scales. Also, both parameters can act in the same direction in their biological effects. For example, the environmental persistence of an exhaled infectious virus particle, associated with a small (<5 micron) or a larger (100 micron) droplet (29), increases under laboratory conditions with lower temperatures and humidity for both influenza (30), SARS (31) and SARS-CoV-2 (32). Although prolonged environmental survival at lower temperatures applies to most of the viruses that have been studied (33), a less humid environment shortens the survival for most upper airway infections such as the rhino-and adeno-viruses that cause the common cold (34). Importantly, however, these viruses lack an envelope and are not seasonally defined (35). In contrast, enveloped viruses, including measles, variola and varicella are infections reaching the lower airways, prevalent during the cooler seasons and predominantly or substantially airborne. It seems plausible that some viruses have adapted to this phase in the seasonal cycle and their concomitant features of temperature and humidity, ruling out the identification of a single or dominant driver of transmission. Our results point to SARS-CoV-2 as belonging to this cluster of viruses.

This membership suggests the importance of airborne transmission even though it has remained controversial so far. Low humidity levels can reduce the size of bio-aerosols by evaporation when these cool-weather viruses are exhaled (36). “Droplet nuclei” smaller than 5 microns in size, can by-pass the nasal zone defenses and enter the deeper parts of the lungs. SARS and SARS-CoV-2 specifically target the ACE-2 protein for cell entry (37) where this protein is most abundant in alveolar tissue (38). During the cooler winter months in higher latitudes, indoor heating generates a micro-climate with low levels of RH, a parameter that better reflects the drying power of air than AH. However, both indoor AH and RH in heated accommodations correspond well with the routinely measured outdoor AH in colder climates (16), and therefore, to the gridded climatology used here for this parameter. The permissive role of low temperatures for the transmission of SARS-CoV-2 appears to contradict the warm-weather related transmission of the second wave in the US for July-August 2020 (not shown). The cooled indoor micro-climate during these months would accommodate transmission, similar to the many outbreaks observed in the meat-processing plants in the US (39), South America and Europe (40), where cooled air is mechanically re-ventilated. The positive correlations we found for the summer months (e.g. Fig 2) during the seasonal low of cases may too reflect community transmission in public temperature-controlled buildings such as care homes (40, 41). They may also result from social clustering and increased contact due to warm weather activities (42). Indeed, air conditioning in the subtropics was also proposed to explain SARS transmission (31), a situation bearing similarities with influenza (43). Regardless of explanation, these positive associations were less frequent overall, and less robust and weaker when zooming into smaller temporal window sizes. Weak climate forcing has been proposed to account for pronounced seasonal variation in models for the population dynamics of influenza (44). A formulation of forcing by specific humidity similar to that used in epidemiological models of seasonal influenza (14) has been recently applied to COVID-19 based on the observed seasonality of other corona viruses (12, 45). Numerical results suggested that the large fraction of the population lacking immunity would override any such environmental forcing, and this “virgin soil” pandemic was projected to continue unabated, throughout the summer. Weak forcing leading to seasonal dynamics was further proposed (10), and the summer recession interpreted as a vindication of effective intervention. Our results support a strong climatic forcing influencing the transmissibility of COVID-19 despite an ample supply of susceptible hosts.

Public health interventions to curb transmission of COVID-19 have focused on reducing the contact rate between people through social distancing, and on handwashing, decontamination of infected surfaces and face-covering. With the exception of face-covering, these measures have emphasized the significance of short-range transmission, the default assumption on respiratory diseases caused by exhaled droplets (46) with a restricted spatial range (47). The role of smaller exhaled droplets that become airborne as bio-aerosols or “droplet nuclei” for extended periods and may cover long distances in a viable state, has been proposed for COVID-19 but has remained unresolved (48–51). Seasonality and the role of low temperatures and associated low humidity, can be mechanistically linked to viable SARS-CoV-2 aerosols, supporting the significance of the airborne transmission route implicated in other studies (52). This link warrants an emphasis on “air-hygiene” through improved indoor ventilation (54)(53) to more effectively intervene in the unfolding pandemic. It also underscores the need to include appropriate meteorological parameters in the evaluation and planning of pharmacological and behavioural control measures.

## Methods

### Data Collection

Data on Covid-19 daily incidence counts were retrieved from the ECDC dataset (54) for the nationally aggregated data and from the COVID19 Data Hub (55) for the NUTS2-disaggregated incidence data. National and regional boundaries were obtained from shapefiles provided by GISCO services by Eurostat (56). Meteorological data were retrieved from ERA5 reanalysis through API requests to the Copernicus Climate Data Store (CDS) (57). We obtained temperature (at 2m), sea level pressure, total precipitation and dew point temperature data for a 0.5×0.5° grid for the country-wise analysis and at a 0.25×0.25° grid for the rest. From the 2m and the dew point temperature, we derived relative humidity, and used the atmos Python package (58) to calculate absolute humidity.

The Gridded Population of the World (GPWv4), developed by the Center for International Earth Science Information Network (CIESIN) at Columbia University, was obtained from (59) at a 2.5 min resolution. We did not include data from the USA in our analyses given the large geographical extension of this country which includes different climatic zones, and the largely asynchronous implementation of intervention policies across its different states.

### Global Statistical Analysis

Our first attempt to identify plausible effects of meteorological covariates on Covid-19 spread applied a comparative regression analysis. To this end, we focused on the exponential onset of the disease, as it is the epidemic phase that allows for a better comparison between countries or regions, without the confounding effect of intervention policies. We first determined for each of the spatial units (either countries or NUTS2 regions), the day in which 20 or more cumulative cases were officially reported. We then fitted the first-order polynomial function *f* (*t*) = *x*_0_ + *rt* for the next 20 days of log-transformed data. The r parameter can be understood as the exponential growth rate, and is then used to estimate the basic reproduction number (*R*_0_) using the estimated serial interval T for COVID19 of 4.7 days (60) such that *R*_0_ = 1 + *rT* (61). (We note that we are interested here in the relationship between the reproductive number and not in the actual inference of *R*_0_). Once *R*_0_ was obtained for all our spatial units, we filtered our meteorological data to match the same fitting period (with a 10-day negative delay to account for an incubation and reporting lapse) for every spatial unit. To compute a single average of the meteorological variables per regional unit, we computed a weighted average on the basis of the population contribution of each grid cell to the total population of the region. We did so to have an aggregated value that would better represent the impact of these factors on the population transmission of COVID19, as the same variation in weather in a high density urban area is more likely to contribute to a change in population level transmission than that of an unpopulated rural area. We then averaged the daily values of temperature and absolute humidity for each country and computed univariate linear models for each of these variables as predictors of *R*_0_. Given the somewhat arbitrary criteria to select the dates to estimate the *R*_0_ in each country, a sensitivity analysis was run to test the robustness of the regressions to changes in the related parameters. We tested 70 different combinations of two parameters: the total number of days used for the fit (18 to 27), and the threshold of cumulative COVID19 cases used to select the initial day of the fit (15 to 45). We also calculated the weather averages by shifting the selected dates accordingly. Then, a linear model for each of the estimates was fitted for both T and AH. A summary of the distribution of parameter estimates (the regression slope coefficients and the R^2^ of the models) is shown in Fig. S2.

### Bivariate time series analysis with scale dependent correlations

To examine associations between cases and climate factors in more detail, a Scale-dependent Correlation Analysis (SDC) was performed on the daily time series of both COVID-19 incidence and a given meteorological variable. SDC is an optimal method for identifying dynamical couplings in short and noisy time series (see (18, 19)). In general, Spearman correlations between incidence and a meteorological time series assess whether there is a monotonic relation between the variables. SDC analysis was specifically developed to study transitory associations that are local in time at a specified temporal scale corresponding to the size of the time intervals considered (s). The two-way implementation (TW-SDC) is a bivariate method that computes non-parametric Spearman rank correlations between two time series, for different pairs of time intervals along these series. Different window sizes (s) can be used to examine increasingly finer temporal resolution. The results are sensitive to the value of this window size s, with expected significant and highest correlation values at the scale of the transient coupling between variables. Correlation values decrease in magnitude as window size increases and averages are computed over too long a time interval. Values can also decrease and become non-significant for small windows when correlations are spurious. Here, the method was applied for windows of different length (from s=75 to 14 days), and despite a weekly cycle shows up in some cases for small s, results removing this cycle were robust. We therefore did not remove this cycle.

Results are displayed in a figure with the following subplots: (1) the two time series to the left and top of the matrix of correlation values, respectively; (2) the matrix or grid of correlation values itself in the center, with significant correlations colored in blue when positive and in red when negative, with rows and columns corresponding to the temporal localization of the moving window along the time series on the left and top respectively; (3) a time series at the bottom, below this grid, with the highest significant correlations for a given time (vertically, and therefore, for the variable that acts as the driver, here the meteorological time series). To read the results, one starts at the diagonal and moves vertically down from it to identify a given lag for which significant correlations are found (the closest to the main diagonal). In some of the SDC figures, the time intervals with high local correlations are highlighted with boxes. These intervals alternate with other ones left blank for which no significant correlation is found. All colored areas correspond to significance levels of at least p<0.05. Significance is assessed with a non-parametric randomization test. (see (18) for further details, and for examples illustrating the method).

Time series approaches do not present the typical problems of comparative spatial regression studies, which might be biased by uncontrollable confounding effects across spatial locations. Our approach focuses on analyzing temporal associations in one such location at a time, and comparing across these locations the patterns of association themselves, including their timing (e.g. in the waning or rising phase of epidemic waves). This allows comparison of results among distinct regions, despite differences in control measures and disease epidemic state.

One reason why couplings between two variables in ecological or epidemiological systems may be transient couplings is the existence of thresholds above or below which responses to forcing are weak or absent. To interrogate our analyses for the existence of critical thresholds/ranges for optimal transmission of the virus (inferred from COVID-19 disease outcomes), we pooled all negative and positive significant correlations performed at a size s=21 days between each meteorological variable and COVID-19 cases. We then computed the proportion of those negative correlations among all possible correlations for a given fragment size obtained for each bin of T or AH values and plotted their distribution (e.g. Fig.5, Fig. S8 and Fig. S9), for all individual regions in Italy.

## Data Availability

All data used is public and obtained from public repositories, as indicated in the manuscript

## Acknowledgments

The authors acknowledge the support of Albert Navarro in the development of SDC apps.

## Funding

AF acknowledges the financial support of HELICAL as part of the European Union’s Horizon 2020 research and innovation programme under the Marie Sklodowska-Curie Grant Agreement No.81354. XR acknowledges support from the Spanish Ministry of Science and Innovation through the ‘Centro de Excelencia Severo Ochoa 2019 2023’ Program (CEX2018 000806S), and support from the Generalitat de Catalunya through the CERCA Program. A.SJ. was supported by a fellowship from “la Caixa” Foundation, Spain (ID 100010434, fellowship code LCF/BQ/DR19/11740017);

## Author contributions

Author contributions: **AF**: Methodology, Software, Formal Analysis, Data Curation, Visualization, Writing - Review & Editing. **MB**: Writing - Original Draft, Writing - Review & Editing. **ASJ**: Software, Formal Analysis, Data Curation. **MP**: Writing - Original Draft, Writing - Review & Editing **XR**: Conceptualization, Methodology, Supervision, Software, Writing - Original Draft, Writing - Review Editing.

## Competing interests

The authors declare no competing interests.

## Data and materials availability

All data used in this study is publicly available at the source websites listed. Code for the analysis will be publicly available in an open repository at time of publication.

## Supplementary Data

**Fig. S 1:**
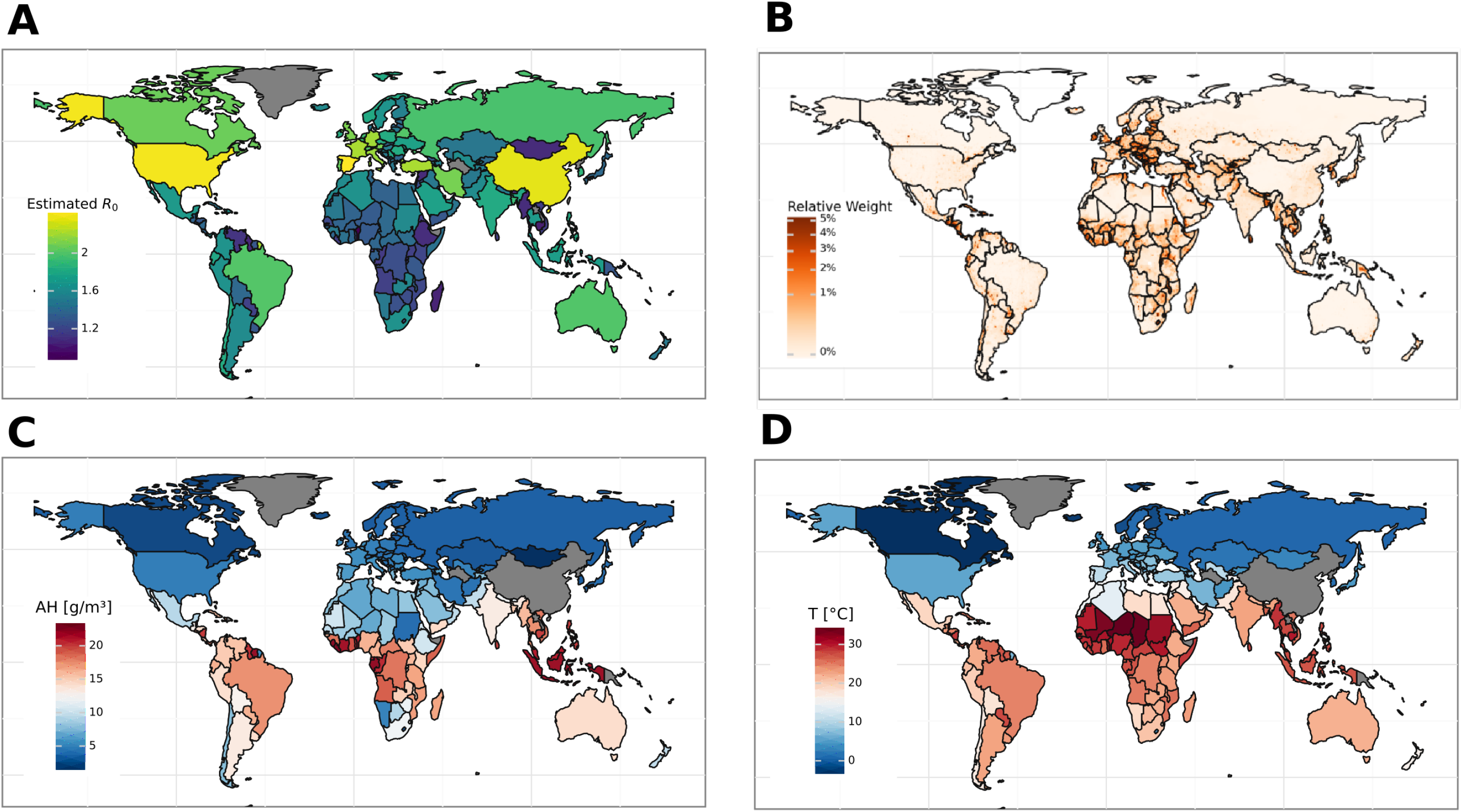
(A) Geographical distribution for *R*_0_ when it is estimated using the period of 20 days after the 100th COVID19 case (for comparison with Fig. 1A. (B) Geographical distribution of relative weights given to each cell in a 1×1° grid. Weights represent the fraction of the population living in each cell for every nation. Maps in C and D correspond to the averages of AH and T +/-10 days before/after the notification day of the 20th COVID19 case in each country, respectively.

**Fig. S 2:**
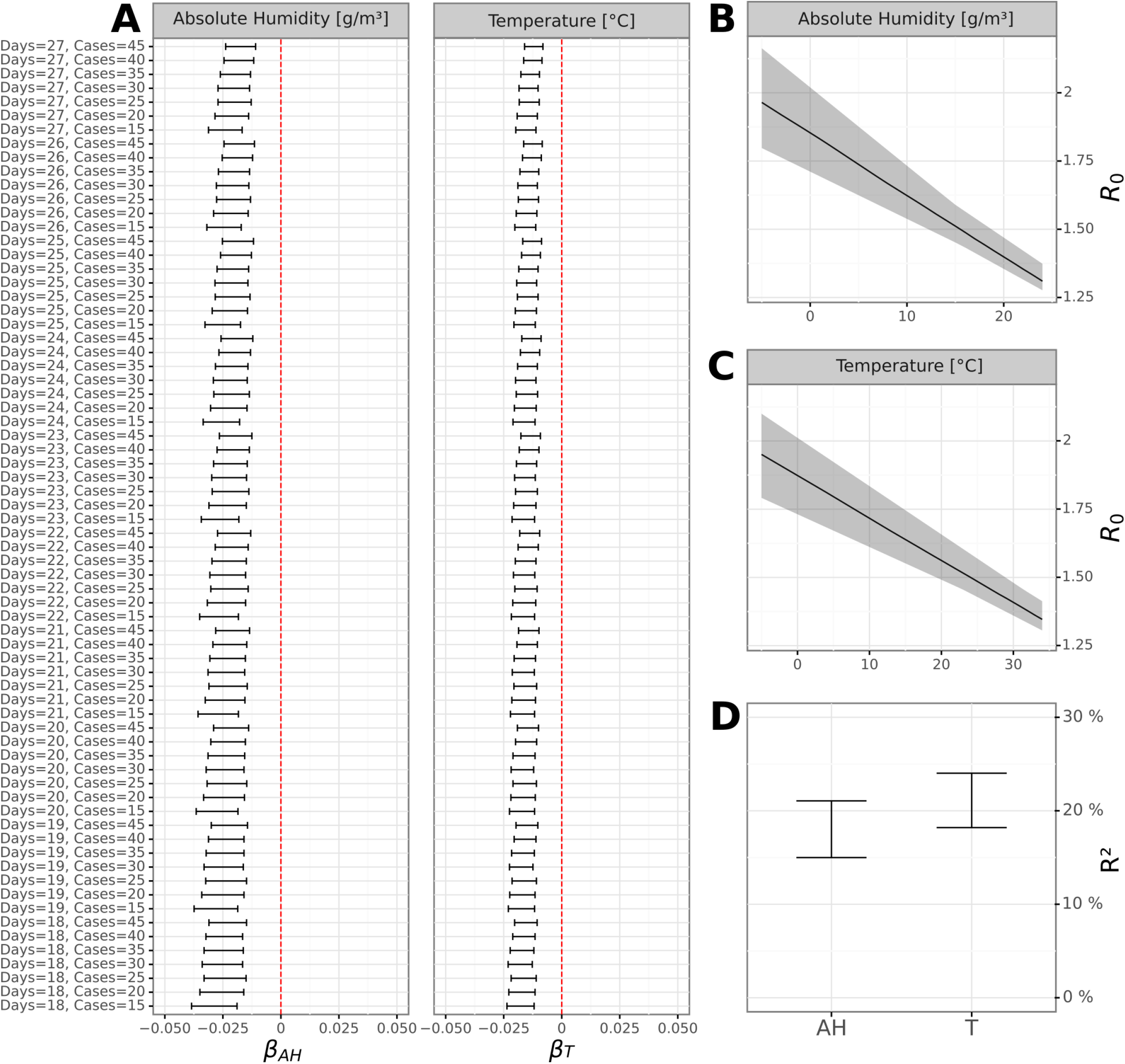
Sensitivity tests for the OLS regression models fitted in Fig. 1B,C. A) 95% CI of the estimated slopes of the OLS regressions (*β*_*AH*_ and *β*_*T*_) for different values of the number of days (18 to 27) and initial number of cases (15 to 45) used for estimating the R0 (see Methods). B and C show the distribution of all the regression lines obtained when considering the variation in days and cases shown in A, for AH and T, respectively. Solid line is the median regression line and the shaded interval corresponds to the area occupied by the most extreme regression lines among the 70 sets of parameters tested. (D) Full range of R^2^ estimates obtained as a result of the sensitivity analysis. Results support the robustness and stability of results in Fig. 1B,C.

**Fig. S 3:**
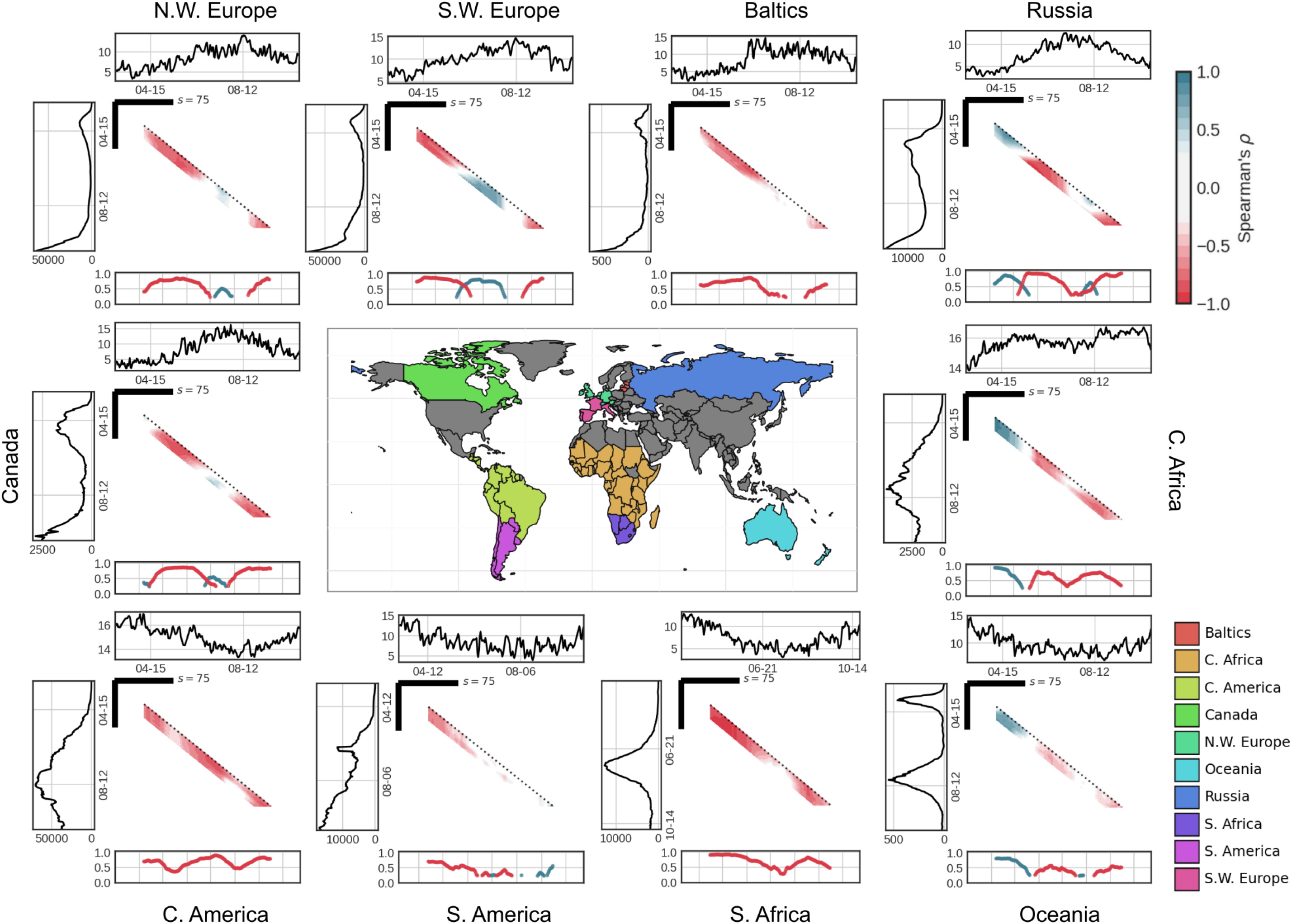
Idem as in Fig. 2 but for AH (g/m3).

**Fig. S 4:**
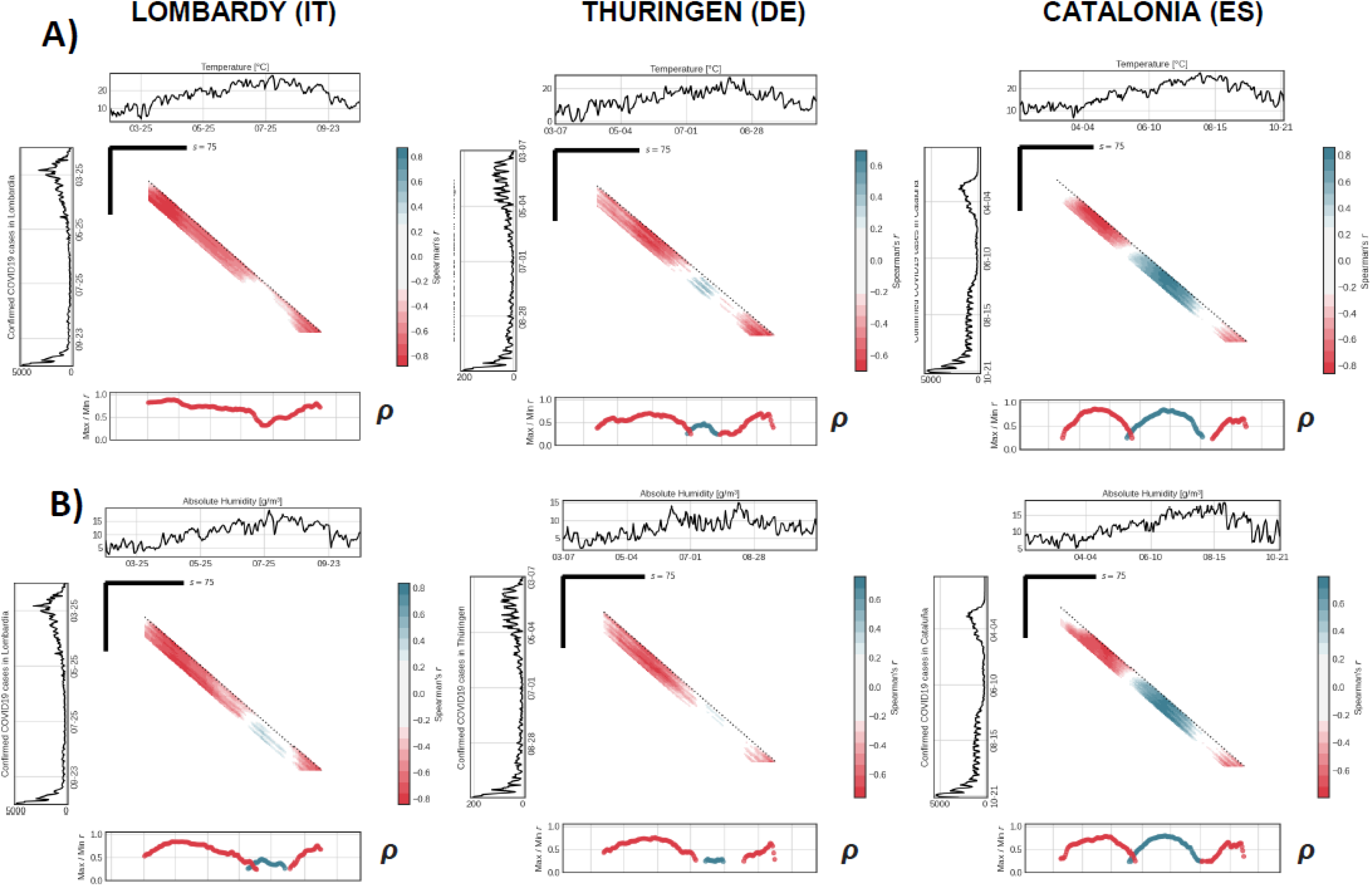
Two-way SDC plots (s=75 and with lags from 0 to 21 days) for the aggregated COVID19 daily new cases time series of three regions in highly affected countries in Europe, namely Italy (Lombardia); Germany (Thüringen) and Spain (Catalonia), and their daily population density-weighted averages of (A) T and (B) AH from March to October 2020. In the figures, each cell of the grid is coloured according to the Spearman correlation coefficient for two s-day centred windows for each of the time series, with distances from the diagonal indicating the temporal lag across the two time series compared. Panels above and left display the time series and panels below show the maximum correlation coefficient obtained for each data point in time. Only those comparisons with a significant p-value at an α=0.05 from a non-parametric randomized test are shown.

**Fig. S 5:**
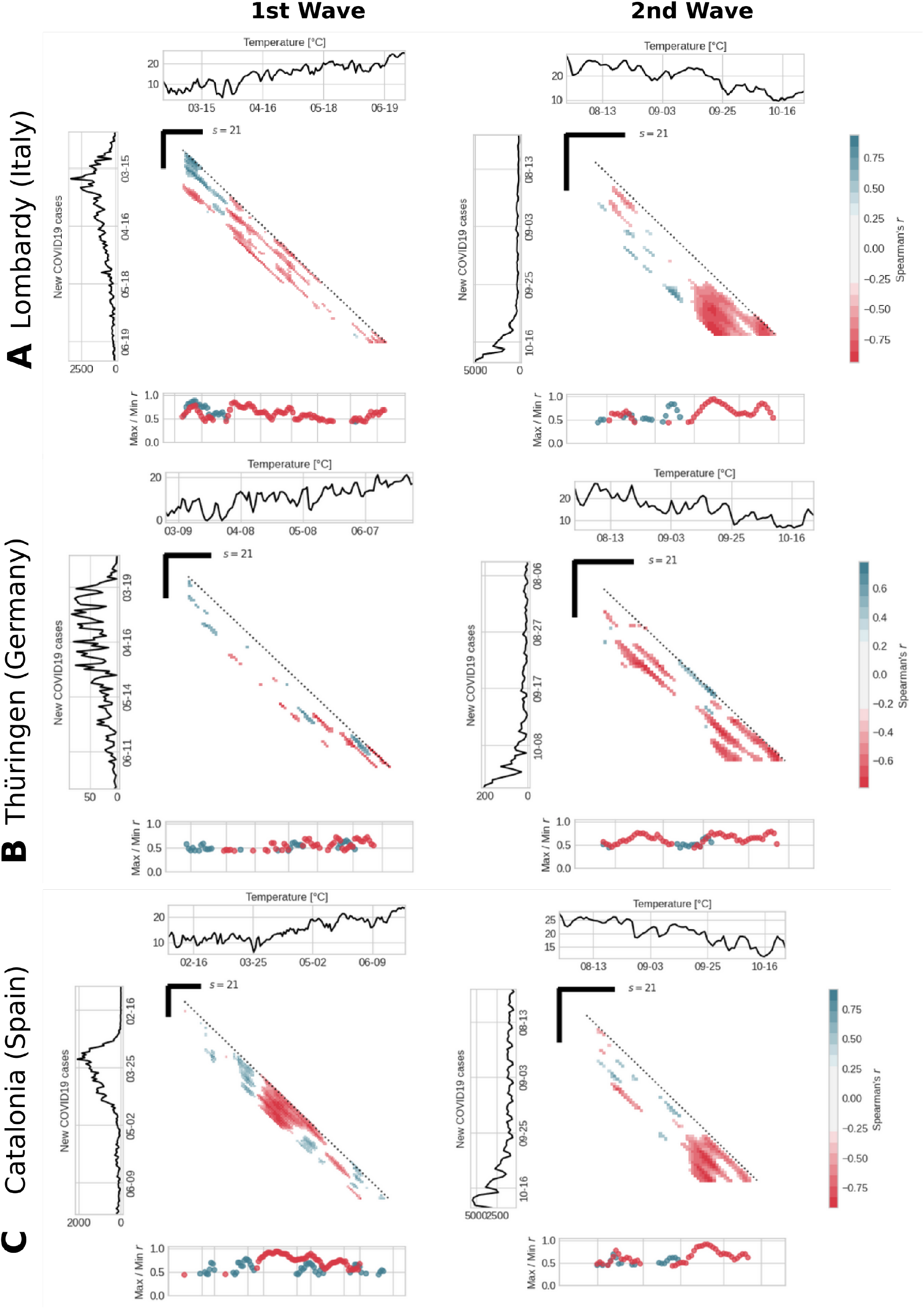
SDC analyses performed at a high temporospatial resolution. The time series are broken down in two intervals: from March to June 2020 to represent the first wave (left), and from August to October 2020 to represent the second wave (right). Fragment sizes used in these analyses are s=21 days, which allows to identify multiple short-term transient couplings between the COVID19 incidence and meteorological factors. Three subnational regions are shown (A-Lombardy in Italy, B-Thüringen in Germany and, C-Catalonia in Spain).

**Fig. S 6:**
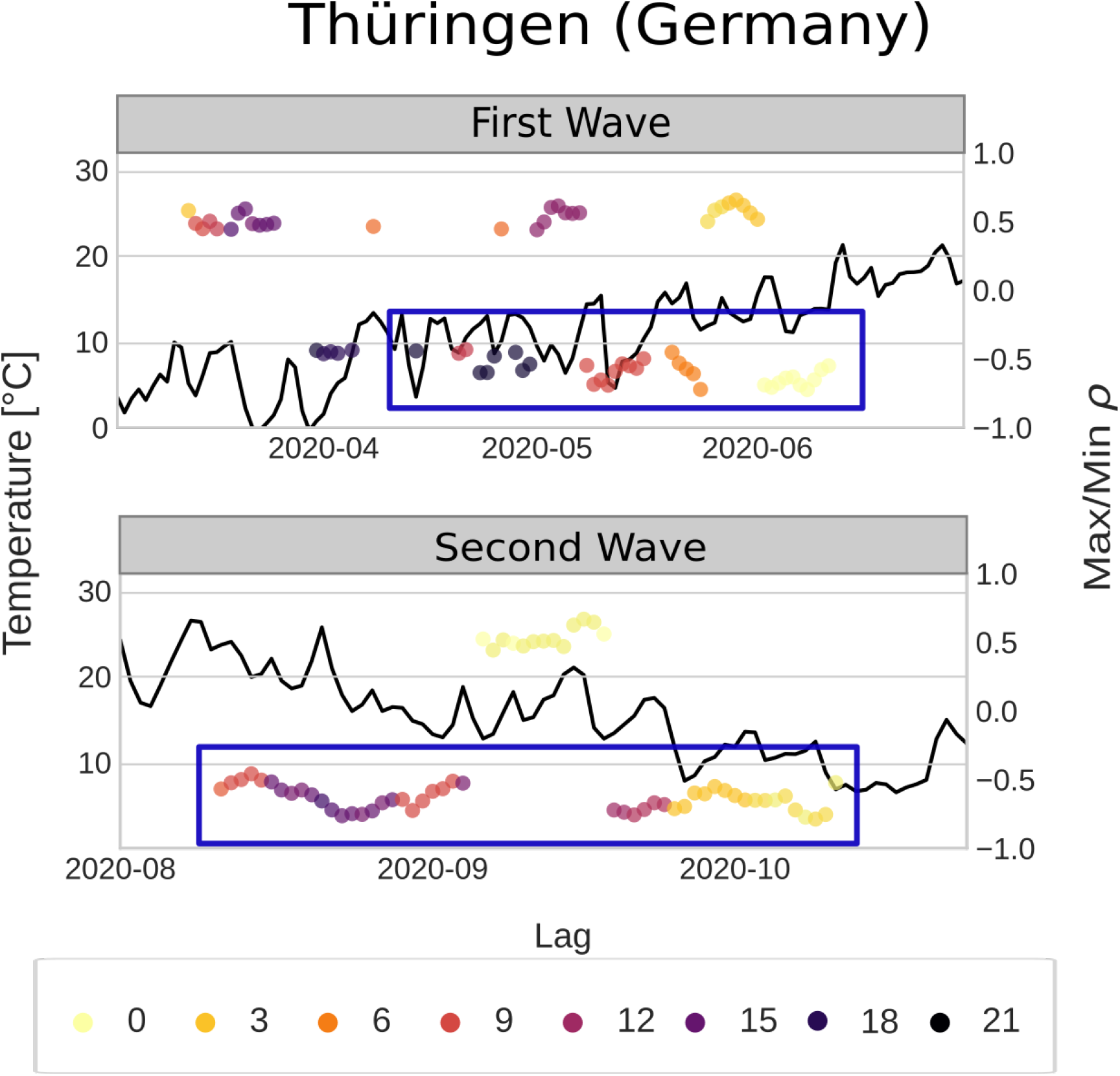
As in Fig. 3B,C but for Thüringen (Germany).

**Fig. S 7:**
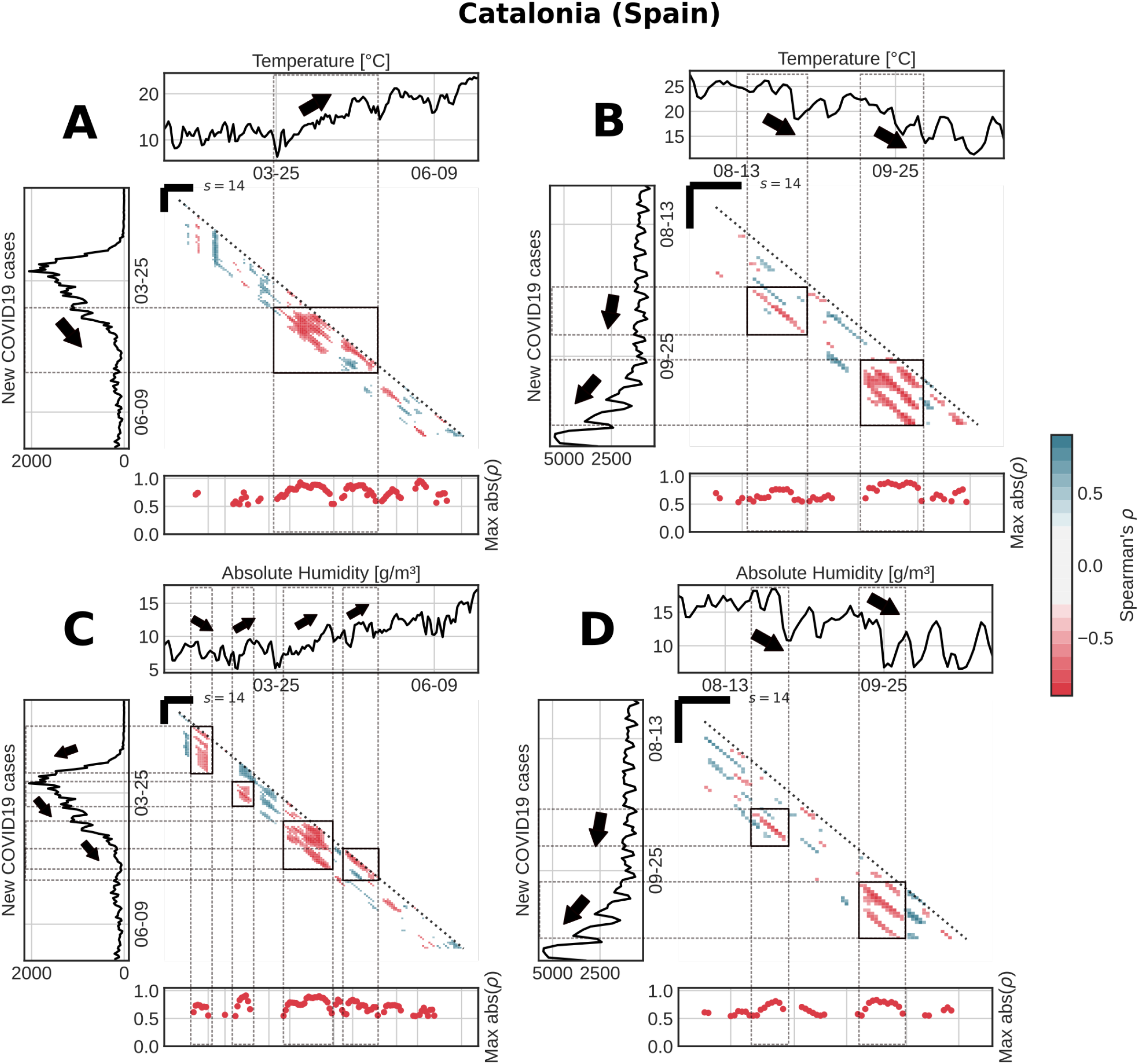
As in Fig. 4 but for Catalonia (Spain).

**Fig. S 8:**
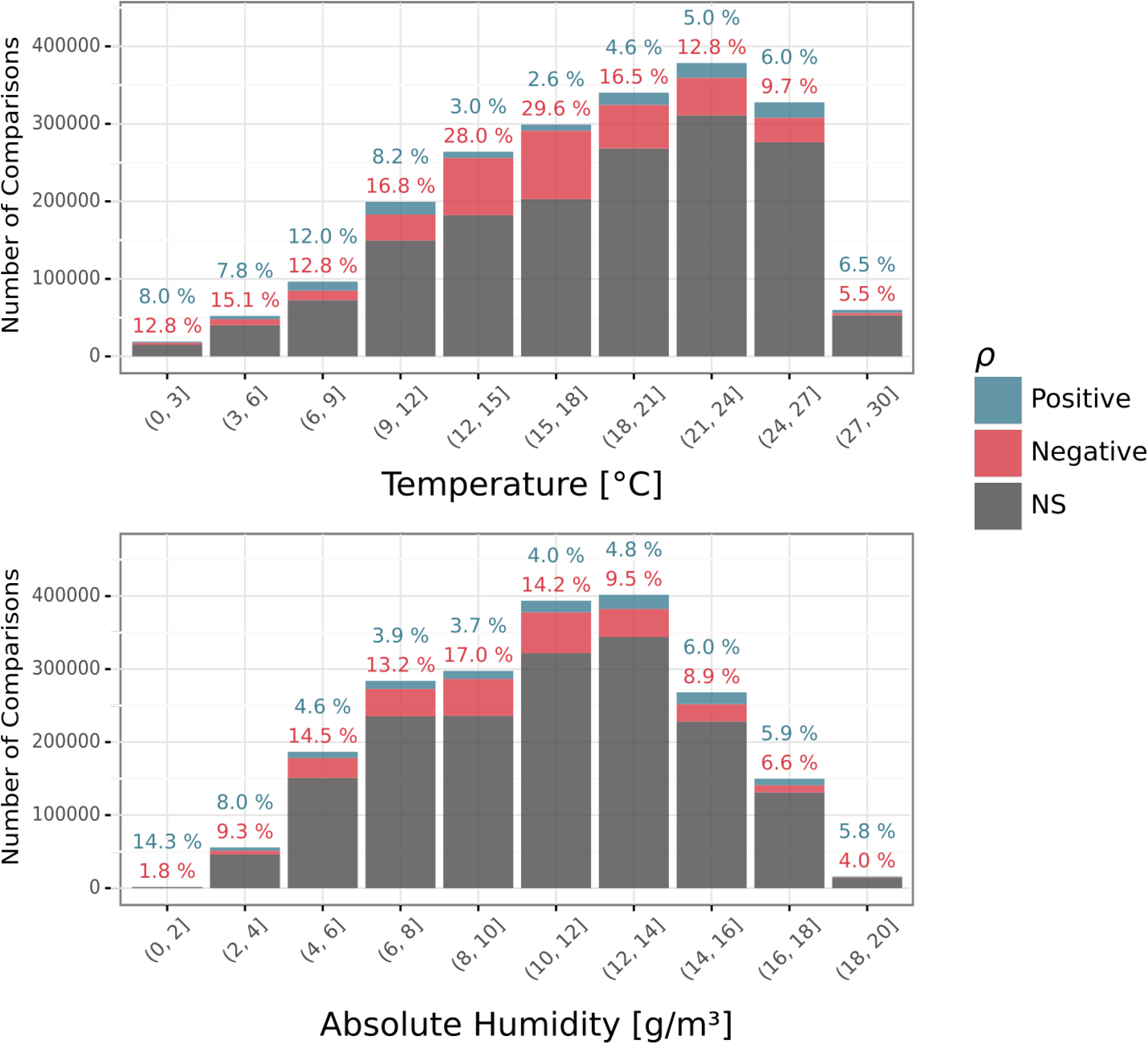
Proportion of significant correlations of (A) T and (B) AH with COVID19 cases. f SDC analysis (s=21 days) is computed with respect to all the comparisons made containing each range of values of the forcing variable, for time series covering all italian regions (n=20). Colours denote negative (red), positive (blue) or non-significant (dark gray) Spearman rank correlations (see Methods).

**Fig. S 9:**
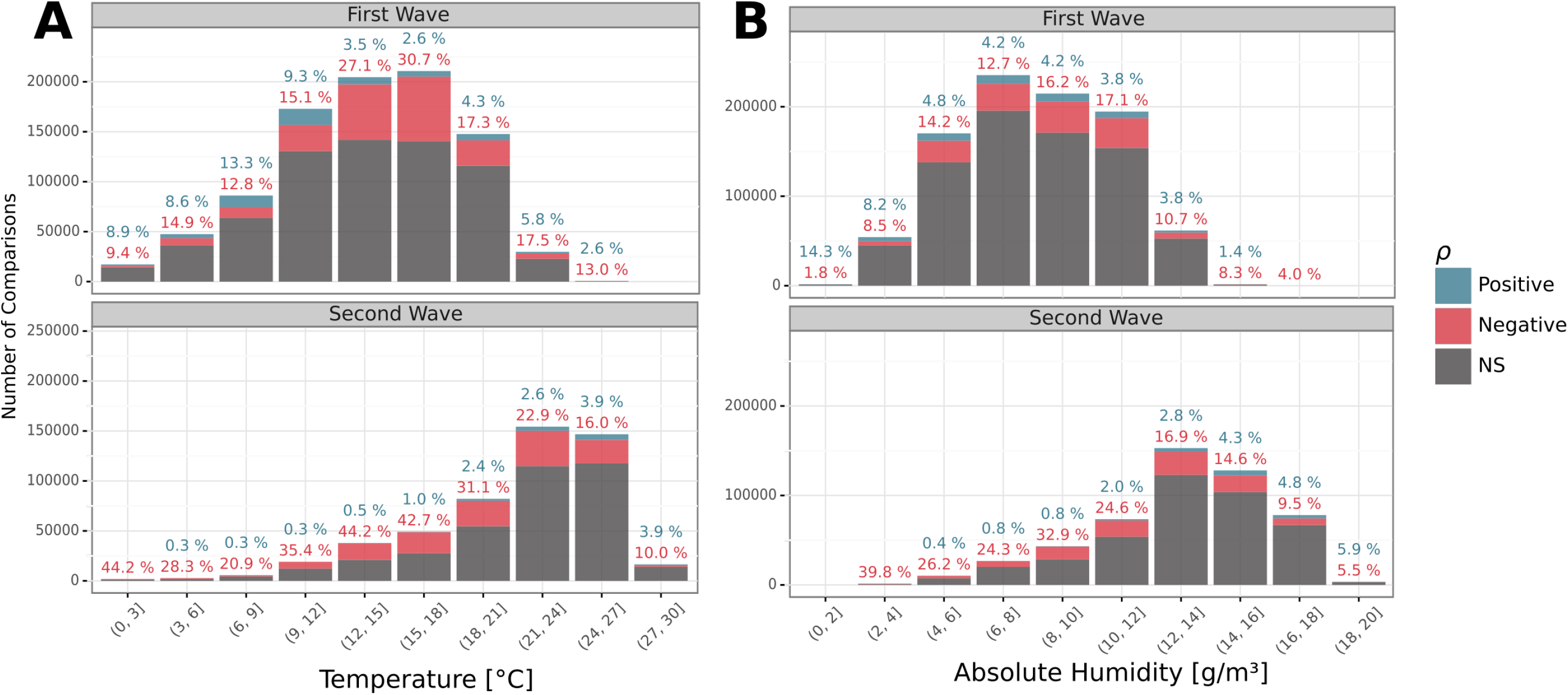
Same as Fig. S8 but separating between the (A) T and; (B) AH, for the first (top) and second (bottom) pandemic waves.

**Fig. S 10:**
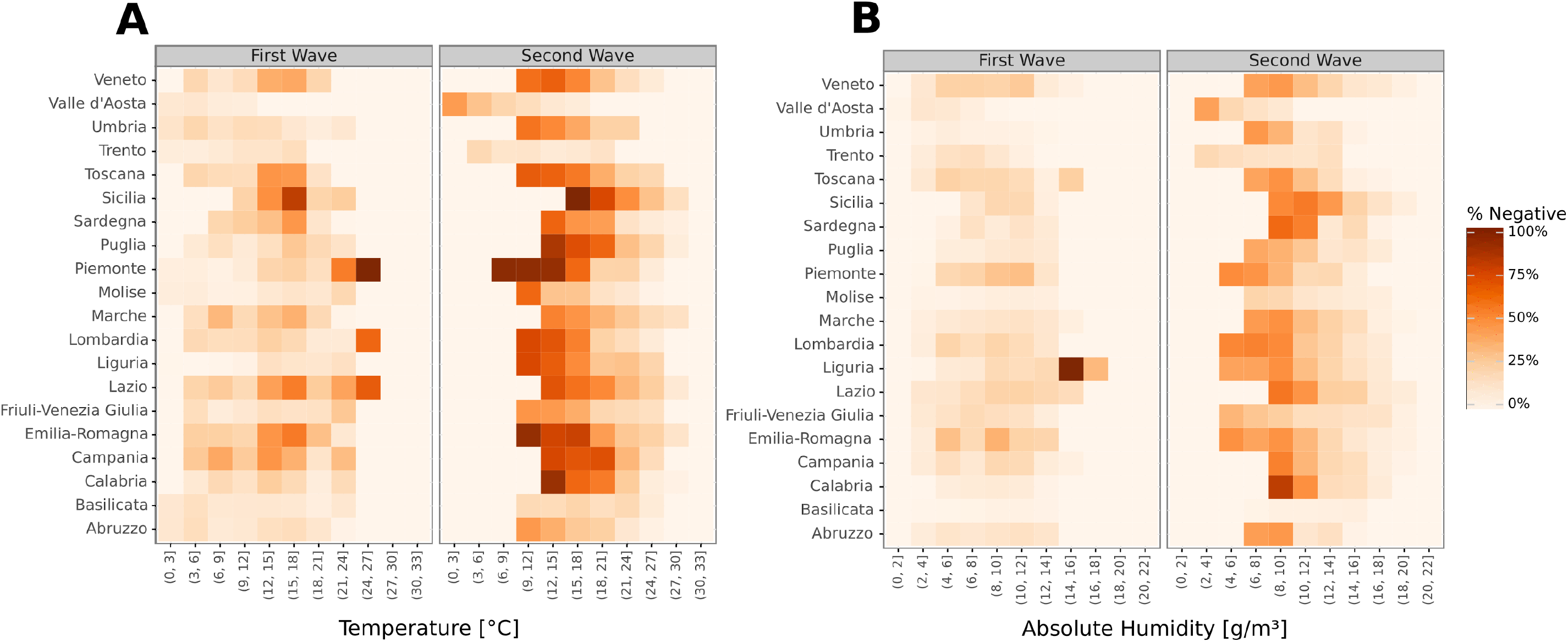
Heatmap representation for all separate regions in Italy, of the proportion of significant negative relationships obtained with a SDC analysis for s=21. Results are shown for the range of values covered by each meteorological variable analyzed (T, AH) and for each of the Italian regions studied. (A) T; (B) AH. Colour scale denotes the percentage of significant negative monotonic relationships in temperate conditions. A and B are split between the first and second waves, respectively.

**Fig. S 11:**
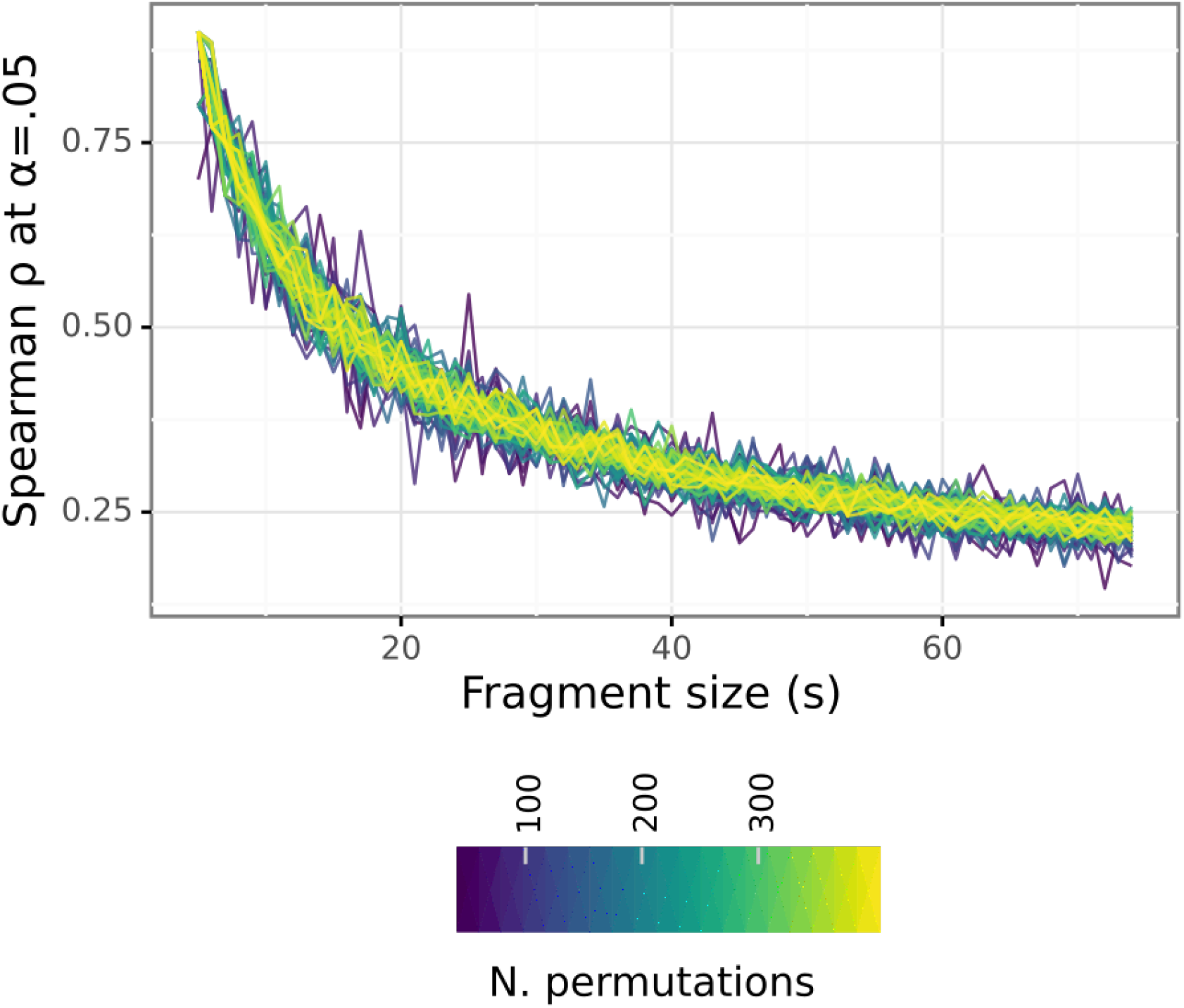
Line plot showing the smallest value of Spearman ρ which yields a significant estimate (at α=.05) as a function of the fragment size s when running a non-parametric permutation test. Here, the permutation test computes the Spearman coefficient ρ between two white noise time series, with each permutation randomly shifting the indices of the series, breaking their temporal shape. This permutation test enables to estimate the probability of finding spurious significant correlations, and can thus be used as a non-parametric test of significance for pairs of time series of interest. The threshold decays rapidly as the fragment size grows, with high ρ values being rarer the longer the time series compared. As indicated by the colour of each line, estimation of the significance threshold becomes more stable as the number of permutations used to estimate it increases.

## Notes

### Competing Interest Statement

The authors have declared no competing interest.

### Funding Statement

AF acknowledges the financial support of HELICAL as part of the European Union Horizon 2020 research and innovation programme under the Marie Sklodowska-Curie Grant Agreement No.81354. XR acknowledges support from the Spanish Ministry of Science and Innovation through the Centro de Excelencia Severo Ochoa 2019 2023
Program (CEX2018 000806S) and support from the Generalitat de Catalunya through the CERCA Program. A.SJ. was supported by a fellowship from la Caixa Foundation, Spain
(ID 100010434, fellowship code LCF/BQ/DR19/11740017).

### Author Declarations

The current study requires no ethics committee aproval. Data used are public fully anonymized records at an aggegate scale of regions in countries.

